# Reduced inflammatory responses to SARS-CoV-2 infection in children presenting to hospital with COVID19 in China

**DOI:** 10.1101/2020.07.02.20145110

**Authors:** Guoqing Qian, Yong Zhang, Yang Xu, Weihua Hu, Ian P Hall, Jiang Yue, Hongyun Lu, Liemin Ruan, Maoqing Ye, Jin Mei

## Abstract

**Background:** Infection with severe acute respiratory syndrome coronavirus 2 (SARS-CoV-2) in children is associated with better outcomes than in adults. The inflammatory response to COVID-19 infection in children remains poorly characterised.

**Methods:** We retrospectively analysed the medical records of 127 laboratory-confirmed COVID-19 patients aged 1 month to 16 years from Wuhan and Jingzhou of Hubei Province. Patients presented between January 25^th^ and March 24^th^ 2020. Information on clinical features, laboratory results, plasma cytokines/chemokines and lymphocyte subsets were analysed.

**Findings:** Children admitted to hospital with COVID-19 were more likely to be male (67.7%) and the median age was 7.3 [IQR 4.9] years. All but one patient with severe disease was aged under 2 and the majority (5/7) had significant co-morbidities. Despite 53% having viral pneumonia on CT scanning only 2 patients had low lymphocyte counts and no differences were observed in the levels of plasma proinflammatory cytokines, including interleukin (IL)-2, IL-4, IL-6, tumour necrosis factor (TNF)-*α*, and interferon (IFN)-*γ* between patients with mild, moderate or severe disease.

**Interpretations:** We demonstrated that the immune responses of children to COVID-19 infection is significantly different from that seen in adults. Our evidence suggests that SARS-CoV-2 does not trigger a robust inflammatory response or ‘cytokine storm’ in children with COVID-19, and this may underlie the generally better outcomes seen in children with this disease. These data also imply anti-cytokine therapies may not be effective in children with moderate COVID-19.

**Funding:** This study was funded by National Natural Foundation of China (No. 81970653).

**Research in context:** *Evidence before this study:* We searched PubMed without language restriction for studies published until June 25, 2020, using the search terms “SARS-CoV-2” or “novel coronavirus” or “COVID-19” and “immune responses” or “innate immunity” or “cytokine” or “subset of lymphocyte” and “children” or “adolescent”. Previously published research describes that severe and fatal cases in children are relatively rare. However, the inflammatory responses to COVID-19 infection in children remains poorly characterised.

*Added value of this study:* We analysed data from 127 laboratory-confirmed COVID-19 patients aged 1 month to 16 years in Hubei province to explore the immune responses to SARS-CoV-2 infection presenting to hospital with COVID-19. Cell numbers of CD3+, CD4+, CD8+ and natural killer T cells were within mostly normal limits even in more severe cases, and the levels of immunoglobulins, and proinflammatory cytokines, including interleukin (IL)-2, IL-4, IL-6, tumour necrosis factor (TNF)-α, and interferon (IFN)-γ were not generally elevated regardless of disease severity.

*Implications of all the available evidence:* The immune response to SARS-CoV-2 infection of children is significantly different from that seen in adults. The inflammatory responses seen even in children with viral pneumonia on CT are relatively mild and do not trigger the “cytokine storm” seen in some adults with COVID-19. This implies anti-cytokine therapies may not be effective in children with COVID-19.

## Introduction

In December 2019, a novel coronavirus was identified in Wuhan China named by the World Health Organization (WHO) severe acute respiratory syndrome coronavirus 2 (SARS-CoV-2). SARS-CoV-2 has 88% sequence homology with SARS-like coronavirus.^1,2^ The WHO declared novel coronavirus disease 2019 (COVID-19) after infection by SARS-CoV-2 a public health emergency of international concern.^3^ By 25 June 2020, there had been 9.5 million cases of COVID-19 and 485,632 deaths globally.^4^

The immune system plays a crucial role in mediating the response to SARS-CoV-2 infection, with three major classes of pattern recognitions receptors (PRRs): toll-like receptors, RIG-I-like receptors (RLRs), and NOD-like receptors (NLRs).^5,6^ These can all trigger downstream signalling effectors, such as Nuclear Factor-*κ*B and IRF3/7, which in turn stimulate anti-viral effector systems including those mediated through CD4+, CD8+, Natural Killer (NK) cells, and macrophages.^7^ Increased release and transcription of proinflammatory cytokines, which may lead to a “cytokine storm”, result in elevated plasma levels of interleukin (IL)-2, IL-6, IL-7, IL-10, IL-1*β*, tumour necrosis factor (TNF)-*α*, and interferon (IFN)-*γ* in some severe or critically unwell subjects. In adults, cytokine responses are correlated with disease severity and poor prognosis in COVID-19 patients.^8,9^ Moreover, pathological findings of COVID-19 biopsy samples have shown interstitial mononuclear inflammatory infiltrates in lung tissues.^10^

However, much less is known about the inflammatory responses in children with COVID-19. Children in general have much milder disease, although a rare subgroup has been identified with a Kawasaki like syndrome termed paediatric inflammatory multisystem syndrome temporally associated with SARS-CoV-2 (PIMS-TS).^11,12^ In this study, we investigated the immune response of children with COVID-19, by measuring immunoglobin, proinflammatory cytokines, and lymphocyte subsets, and in particular T-cell responses during the early and late stage of SARS-CoV-2 infection, to provide insight into the role of immune response in children with COVID-19.

## Methods

### Study population and clinical samples

We performed a retrospective cohort to study the immune response of 127 COVID-19 children subjects aged 1 month to 16 years, who were admitted into the Wuhan Children’s Hospital and Jingzhou First People’s Hospital during January 25^th^ and March 24^th^, 2020. The diagnosis of COVID-19 was based on the guidelines issued under the New Coronavirus Pneumonia Prevention and Control Program (4^th^ edition) published by the National Health Commission of China,^13^ all included cases were confirmed by real-time reverse transcriptase polymerase-chain-reaction (RT-PCR) using nasal swab specimens.

This study was conducted in accordance with the Declaration of Helsinki and was reviewed and approved by the Medical Ethical Committees (2020-R120). Due to the urgent need to collect data on this emerging infectious disease, the requirement for written informed consent was waived.

### COVID-19 clinical classification

Cases were classified into mild, moderate, severe and severe/critical cases following the guidelines issued by the National Health Commission of China.^13^

1. Mild Mild clinical symptoms without lung infiltration on chest X ray or chest computed tomography (CT) scan.
2. Moderate Fever, cough and other symptoms present alongside lung infiltrations on chest X ray or chest CT scan.
3. Severe (children-specific definition) The subject was classified as severe if at least one of the following diagnosis conditions was met:
  a. Shortness of breath as defined by age (<2 months, RR ≥ 60 per min; 2-12 months, RR ≥ 50 per min; 1-5 years, RR ≥ 40 per min;> 5 years, RR ≥ 30 per min).
  b. Oxygen saturations whilst breathing room air and at rest ≤93%;
  c. Respiratory distress, cyanosis, or intermittent apnoea;
  d. Drowsiness and convulsions;
  e. Refusal to eat or feeding difficulties, signs of dehydration.
4. Critical One of the follow conditions has to be met:
  a. Respiratory failure requiring mechanical ventilation
  b. Cardiovascular Shock.
  c. Patients with other organ dysfunction needing intensive care unit treatment.

### Cytokine assays

The levels of plasma cytokines or chemokines (IL-2, IL-4, IL-6, IL-10, TNF-*α*, and IFN-*γ*) was quantified by enzyme linked immunosorbent assay (ELISA) kits. All assays were performed according to the manufacturer’s instructions.

### Flow Cytometric analysis

Peripheral blood mononuclear cells (PBMCs) from 127 patients were obtained and processed in the Clinical Lab of Wuhan Children Hospital. The PBMCs were harvested by centrifugation and stained using a commercially available kit (Hangzhou Cellgene Biotech Co. Ltd). Cell numbers of total T, CD4+ T, CD8+ T, B cells and NK cells were analysed by flow cytometry on and LSR Fortessa Cell Analyzer (BD Biosciences) and data processed using the FolwJo software.

### Statistical analysis

Continuous variables were described using means and standard deviations (SD) or median (IQR). Comparisons across groups were performed with independent t-tests for normally distributed data or by Mann-Whitney U tests. Chi-square tests and Fisher’s exact test were applied for categorical variables as appropriate. Univariate and multivariate logistic regression analysis was performed to explore the relationship between each variable and the risk of severity of COVID-19. All analyses were analysed using IBM SPSS statistics version 26.0.

### Role of the funding source

This study was funded by National Natural Foundation of China (No. 81970653). The funder had no role in study design, data collection, analysis, and interpretation or writing of this research.

## Results

### Clinical characteristics

A total of 127 children with COVID-19 were admitted to hospital, 67.72% (86/127) were male, and the median age was 7.3 [IQR 4.9] years (Table 1). The subjects were classified into mild, moderate, and severe/critical groups following the guidelines of the National Health Commission of China. Eighty-one children were diagnosed as mild type COVID-19, including one 4-year-old boy with acute lymphoblastic leukaemia (ALL). There were 39 moderate type COVID-19 subjects, including one 1-month-old boy with an atrial septal defect and another 1-month-old boy with intracranial haemorrhage. Severe/critical type (n=7) included 2 fatalities, one in a patient with intussusception and multiorgan failure, and one in a patient with an intracranial malignant tumour. 3 of the other 5 severe/critical patients also had comorbidities, including at the time of initial presentation one with ALL on maintenance chemotherapy, two with atrial septal defects. The other two were otherwise healthy at initial presentation.

**Table 1.** Demographic, baseline features of 127 children or adolescence with COVID-19. Values are numbers, mean±SD (standard deviation) or medians (interquartile ranges) unless stated otherwise.

| **Table 1. Demographic, baseline features of 127 children or adolescence with COVID-19.**<br>Values are numbers, mean±SD (standard deviation) or medians (interquartile ranges) unless stated otherwise. | | | | |
| --- | --- | --- | --- | --- |
|  | **Total** | **Mild** | **Moderate** | **Severe and Critical** |
| **No. of patients** | 127 | 81 | 39 | 7 |
| **Age Mean±SD, yr** | 7.27±4.85 | 8.43±4.46 | 5.73±4.85 | 1.64±2.85 |
| **Distribution, No. of patients** |  |  |  |  |
| 0-12m | 28 | 8 | 15 | 5 |
| 13-24m | 2 | 1 | 0 | 1 |
| >25m, ≤5y | 14 | 12 | 2 | 0 |
| >5y, ≤10y | 45 | 28 | 16 | 1 |
| >10y | 38 | 32 | 6 | 0 |
| **M/F, No.** | 86/39 | 58/23 | 25/14 | 5/2 |
| **Weight (kg) (Mean±SD)** | 35.06±23.9 | 36.88±20.13 | 34.44±28.77 | 11.60±8.08 |
| **Height (cm) (Mean±SD)** | 124.5±36.49 | 135.47±28.01 | 112.24±41.66 | 78.80±27.58 |
| **Vital Signs, median [IQR]** |  |  |  |  |
| Temperature, $^{\circ}$C | 36.5[36.2-36.8] | 36.5[36.3-36.8] | 36.5[36.2-36.8] | 36.7[36.2-37.0] |
| Pulse, per/min | 96[88-110] | 92[87-101] | 100[90-121] | 145[110-160] |
| Respiratory rate, per/min | 22[20-26] | 22[20-24] | 24[20-30] | 26[25-45] |
| **Comorbidity** |  |  |  |  |
| Acute lymphoblastic leukaemia | 2 | 1 | 0 | 1 |
| Intussusception | 1 | 0 | 0 | 1 |
| Atrial septal defect | 3 | 0 | 1 | 2 |
| Intracranial malignant tumour | 1 | 0 | 0 | 1 |
| Intracranial hemorrhage | 1 | 0 | 1 | 0 |
| **Co-infection** |  |  |  |  |
| Mycoplasma IgM (+) | 60 | 42 | 15 | 3 |
| CMV IgM (+) | 4 | 2 | 2 | 0 |
| CMV IgG (+) | 38 | 22 | 14 | 2 |
| EB IgM/VCA (+) | 3 | 3 | 0 | 0 |
| EB IgG/VCA (+) | 49 | 27 | 18 | 4 |
| **Length days of hospitalisation (d) (Mean±SD)** | 15.41±7.86 | 13.35±5.16 | 17.18±7.74 | 30.00±19.58 |
| **Fatal or MV** | 2 | 0 | 0 | 2 |
| COVID-19, coronavirus disease 2019; CMV, cytomegalovirus; EBV, Epstein-Barr virus. MV, mechanical ventilation |  |  |  |  |

All patients with severe/critical disease less than 2 years old (1 month – 16 months) except one 8-year-old boy who developed myocardial damage. CT changes of pneumonia were present in 67/120 patients who underwent CT scans. Except for the two patients who died, all other patients (125) were discharged from the hospital, with an average length of stay of 15.4 days across the whole group 47.24% (60/127) subjects tested positive of mycoplasma IgM, 4 subjects tested positive for cytomegalovirus (CMV), and 3 subjects tested positive for Epstein-Barr virus (EBV) IgM/VCA. Table 2 shows the results of blood tests and CT scan results for the full sample. Lymphopenia (lymphocyte count, <1.0×10^9^ per liter) was only present in 2 patients (1.57%), including one 6-month-old boy (0.37×10^9^/L) with intracranial malignant tumour who died and one 9-year-old boy (0.96×10^9^/L).

**Table 2.** **Clinical characteristics of 127 children or adolescence with COVID-19. Values are mean±SD (standard deviation) or medians (interquartile ranges) unless stated otherwise**.

|  |  |  |  |  |
| --- | --- | --- | --- | --- |
|  | **Total** | **Mild** | **Moderate** | **Severe and Critical** |
| White blood cells (4-10×10<sup>9</sup> cells per L) | 6.61±2.09 | 6.79±2.04 | 6.48±2.04 | 5.33±2.57 |
| Decreased (n) | 9 | 5 | 2 | 2 |
| Neutrophil (2-7×10<sup>9</sup> cells per L) | 2.94±1.98 | 3.18±2.27 | 2.71±1.28 | 1.89±1.80 |
| Decreased (n) | 26 | 15 | 11 | 0 |
| Lymphocytes (1-4×10<sup>9</sup> cells per L) | 3.07±1.52 | 2.92±1.41 | 3.30±1.70 | 3.37±1.81 |
| Decreased (n) | 2 | 0 | 1 | 1 |
| Eosinophil (0.05-0.5×10<sup>9</sup> cells per L) | 0.24±0.38 | 0.28±0.46 | 0.18±0.14 | 0.13±0.17 |
| Decreased (n) | 16 | 9 | 3 | 4 |
| Platelets (100-300×10<sup>9</sup> cells per L) | 298.90±95.87 | 286.42±68.83 | 298.84±116.26 | 314.14±28.47 |
| Decreased (n) | 3 | 0 | 1 | 2 |
| C-reactive protein (<0.75mg/L) | 1.42±1.97 | 0.82±0.19 | 3.88±4.42 | 0.75±0.01 |
| Increased (n) | 76 | 47 | 24 | 4 |
| Procalcitonin (≤0.05ng/ml) | 0.07±0.12 | 0.06±0.13 | 0.07±0.04 | 0.26±0.24 |
| Increased (n) | 38 | 17 | 16 | 5 |
| Fibrinogen (2-4g/L) | 2.17±0.64 | 2.19±0.63 | 2.19±0.72 | 2.07±0.28 |
| Increased (n) | 2 | 1 | 1 | 0 |
| D-dimer (0-0.55mg/L) | 0.44±1.12 | 0.38±1.00 | 0.57±1.38 | 0.15±0.04 |
| Increased (n) | 8 | 5 | 3 | 0 |
| Alanine aminotransferase (21-27 IU/L) | 29.74±58.59 | 20.35±18.93 | 49.80±98.77 | 19.57±22.90 |
| Increased (n) |  | 14 | 15 | 0 |
| Aspartate transferase (15-46 IU/L) | 36.38±61.71 | 29.07±13.89 | 51.63±106.62 | 27.71±6.63 |
| Increased (n) | 55 | 30 | 24 | 1 |
| Creatinine (58-110μmol/L) | 37.38±14.02 | 37.83±14.32 | 36.36±14.00 | 37.5±7.64 |
| Increased (n) | 0 | 0 | 0 | 0 |
| Blood urea nitrogen (3.2-7.1 mmol/L) | 4.20±1.52 | 4.03±1.26 | 4.50±1.89 | 4.75±0.72 |
| Increased (n) | 1 | 0 | 1 | 0 |
| Calcium (2.1-2.55 mmol/L) | 2.47±0.12 | 2.48±0.11 | 2.46±0.13 | 2.45±0.14 |
| Decreased (n) | 24 | 14 | 9 | 1 |
| **Infiltration of lung** |  |  |  |  |
| Pneumonia No. | 67 | 36 | 26 | 5 |
| non-pneumonia No. | 53 | 39 | 12 | 2 |
| No data | 7 | 6 | 1 | 0 |
| No. of lobes affected, median[IQR] | 1[1-2] | 1[1-2.75] | 2[1-2] | 1[1-3.5] |
| Data are mean, unless otherwise indicate. COVID-19, coronavirus disease 2019. |  |  |  |  |

### T-cell immune responses to SARS-CoV-2

Both helper T (Th) cells (CD3+CD4+) and suppressor T cells (CD3+CD8+) counts in children with COVID-19 were within normal levels (Table 3). There were no significant differences in cell counts between the mild, moderate and severe/critical groups with COVID-19. The number of CD3+, CD3+CD4+, CD19+ B cells, and the ratio of CD4+/CD8+ were significantly lower for those aged 10 or above (Table 4).

**Table 3.** T cell number and proinflammatory cytokines of 127 children or adolescence with COVID-19. Values are medians (interquartile ranges) unless stated otherwise.

|  | **Total** | **Mild** | **Moderate** | **Severe and Critical** |
| --- | --- | --- | --- | --- |
| **CD3+T cell proportion (%)** | 68.09[63.61-72.95] | 67.78[63.54-72.83] | 69.60[63.70-73.91] | 70.49[66.87-72.35] |
| **CD3+T cell number** | 2073[1675-2779] | 2020[1609-2503] | 2145[1682-2969] | 2280[1717-2644] |
| **CD3+CD4+T cell proportion (%)** | 35.81[30.78-39.19] | 36.61[31.30-39.76] | 35.69[30.79-37.82] | 35.89[33.24-37.03] |
| **CD3+CD4+T cell number** | 1012[669-1569] | 1011[672-1466] | 1022[677-1667] | 1010[839-1391] |
| **CD3+CD8+T cell proportion (%)** | 27.52[21.84-31.49] | 25.94[21.06-29.73] | 29.11[24.96-32.43] | 28.32[26.32-31.15] |
| **CD3+CD8+T cell number** | 743[556-1022] | 711[490-990] | 784[605-1286] | 1022[692-1220] |
| **Natural killer proportion (%)** | 9.97[5.71-14.59] | 10.40[6.03-15.35] | 8.80[5.44-14.15] | 11.80[6.79-14.62] |
| **Natural killer cell number** | 316[187-470] | 334[188-476] | 308[207-421] | 225[182-417] |
| **Lymphocyte (%)** | 18.15[15.35-22.61] | 19.03[15.60-22.55] | 17.02[14.81-22.74] | 16.50[14.43-19.14] |
| **CD19+B cell number** | 557[372-822] | 553[389-764] | 528[367-878] | 528[450-649] |
| **CD4+/CD8+T ratio** | 1.34[1.09-1.78] | 1.39[1.13-1.81] | 1.34[1.11-1.76] | 1.21[1.16-1.34] |
| **CD4+CD25+T cell proportion (%)** | 4.93[3.64-5.70] | 4.84[3.59-5.67] | 4.70[3.76-6.06] | 5.15[3.74-6.73] |
| **Human IL-2** | 1.43[1.21-1.67] | 1.45[1.26-1.67] | 1.42[1.16-1.65] | 1.54[1.33-1.68] |
| **Human IL-4** | 2.68[2.07-3.33] | 2.71[2.18-3.33] | 2.53[1.94-3.05] | 2.83[2.55-3.42] |
| **Human IL-6** | 3.98[3.04-5.66] | 4.02[3.21-5.30] | 3.95[2.96-8.22] | 3.17[2.68-3.59] |
| **Human IL-10** | 3.50[3.05-4.61] | 3.50[3.11-4.44] | 3.49[2.90-5.66] | 3.26[3.05-4.13] |
| **Human TNF-α** | 1.62[1.26-2.23] | 1.70[1.33-2.27] | 1.55[1.06-1.84] | 1.76[1.5-2.39] |
| **Human IFN-γ** | 2.87[2.23-3.91] | 2.89[2.44-3.91] | 2.84[1.94-3.84] | 2.78[1.85-3.01] |

**Table 4.** **Levels of T cell number and proinflammatory cytokines of 127 children or adolescence with COVID-19 in different age stages. Values are medians (interquartile ranges) unless stated otherwise**.

|  | **Total** | **≤24m** | **>25m, ≤5y** | **>5, ≤10y** | **>10y** |
| --- | --- | --- | --- | --- | --- |
| **CD3+T cell proportion (%)** | 68.09[63.61-72.95] | 66.44[59.76-75.26] | 65.66[59.87-67.23] | 68.97[64.06-72.35] | 69.98[65.31-73.79] |
| **CD3+T cell number** | 2073[1675-2779] | 3234[2329-4403] | 2426[2082-3617] | 1971[1647-2287] | 1868[1554-2267] |
| **CD3+CD4+T cell proportion (%)** | 35.81[30.78-39.19] | 41.81[35.95-46.25] | 30.79[27.96-35.52] | 35.90[30.63-39.18] | 32.10[28.74-36.86] |
| **CD3+CD4+T cell number** | 1012[669-1569] | 2033.5[1757-2456] | 1205[891-1403] | 970[670-1123] | 740[610-904] |
| **CD3+CD8+T cell proportion (%)** | 27.52[21.84-31.49] | 23.46[18.09-27.52] | 27.72[22.54-30.91] | 27.07[22.13-31.12] | 28.80[26.10-31.92] |
| **CD3+CD8+T cell number** | 743[556-1022] | 1100[716-1281] | 1010[788-1590] | 698[481-983] | 707[513-759] |
| **Natural killer proportion (%)** | 9.97[5.71-14.59] | 5.905[4.73-8.54] | 12.21[4.44-18.91] | 11.04[6.05-13.43] | 9.81[6.20-16.52] |
| **Natural killer cell number** | 316[187-470] | 289[220-425] | 437[308-696] | 316[197-407] | 282[145-476] |
| **Lymphocyte (%)** | 18.15[15.35-22.61] | 21.37[17.04-28.14] | 19.96[17.53-27.79] | 17.69[15.09-20.22] | 16.19[14.09-20.69] |
| **CD19+B cell number** | 557[372-822] | 999[725-1687] | 799[534-1204] | 538[358-651] | 408[329-540] |
| **CD4+/CD8+T ratio** | 1.34[1.09-1.78] | 1.87[1.44-2.68] | 1.34[1.22-1.81] | 1.30[1.10-1.61] | 1.14[0.91-1.39] |
| **CD4+CD25+T cell proportion (%)** | 4.93[3.64-5.70] | 4.86[3.53-5.59] | 4.51[3.68-5.02] | 4.65[3.12-5.77] | 5.34[4.38-6.75] |
| **Human IL-2** | 1.43[1.21-1.67] | 1.40[1.27-1.69] | 1.52[1.27-1.65] | 1.47[1.25-1.95] | 1.36[1.18-1.50] |
| **Human IL-4** | 2.68[2.07-3.33] | 2.72[1.78-3.29] | 2.53[2.36-2.94] | 2.58[2.09-3.43] | 2.71[2.16-3.24] |
| **Human IL-6** | 3.98[3.04-5.66] | 4.36[3.51-7.33] | 3.43[2.71-5.13] | 4.14[3.14-6.63] | 3.70[3.06-4.97] |
| **Human IL-10** | 3.50[3.05-4.61] | 4.53[3.47-7.08] | 3.60[3.01-4.35] | 3.71[3.11-5.00] | 3.19[2.79-3.49] |
| **Human TNF-α** | 1.62[1.26-2.23] | 1.80[1.43-2.40] | 1.65[1.28-1.93] | 1.63[1.27-2.68] | 1.50[0.92-1.83] |
| **Human IFN-γ** | 2.87[2.23-3.91] | 3.01[2.57-3.80] | 3.14[2.54-4.06] | 2.78[2.11-4.78] | 2.81[1.85-3.38] |

### Plasma cytokine profiles and immunoglobulins

To investigate the potential role of proinflammatory cytokines/chemokines in SARS-CoV-2 infection, we measured the levels of IL-2, IL-4, IL-6, TNF-*α*, and IFN-*γ* in plasma by ELISA. Interestingly levels were generally low and there were no strong associations with disease severity (Table 3). Similarly, Complement (C3 and C4) and immunoglobulins (IgG, IgA, IgM, and IgE) did not differ between mild, moderate and severe/critical cases (Figure 1). Levels of immunoglobulins (IgG, IgA, IgM and C3) were significantly higher in older children, while IgE and C4 were not associated with age (Figure 2). We observed weak associations between levels of cytokines and frequency of T cell subsets, but apart from a negative correlation between CD3+ counts and TNF-*α* levels none of these survived corrections for multiple testing.

**Figure 1.**
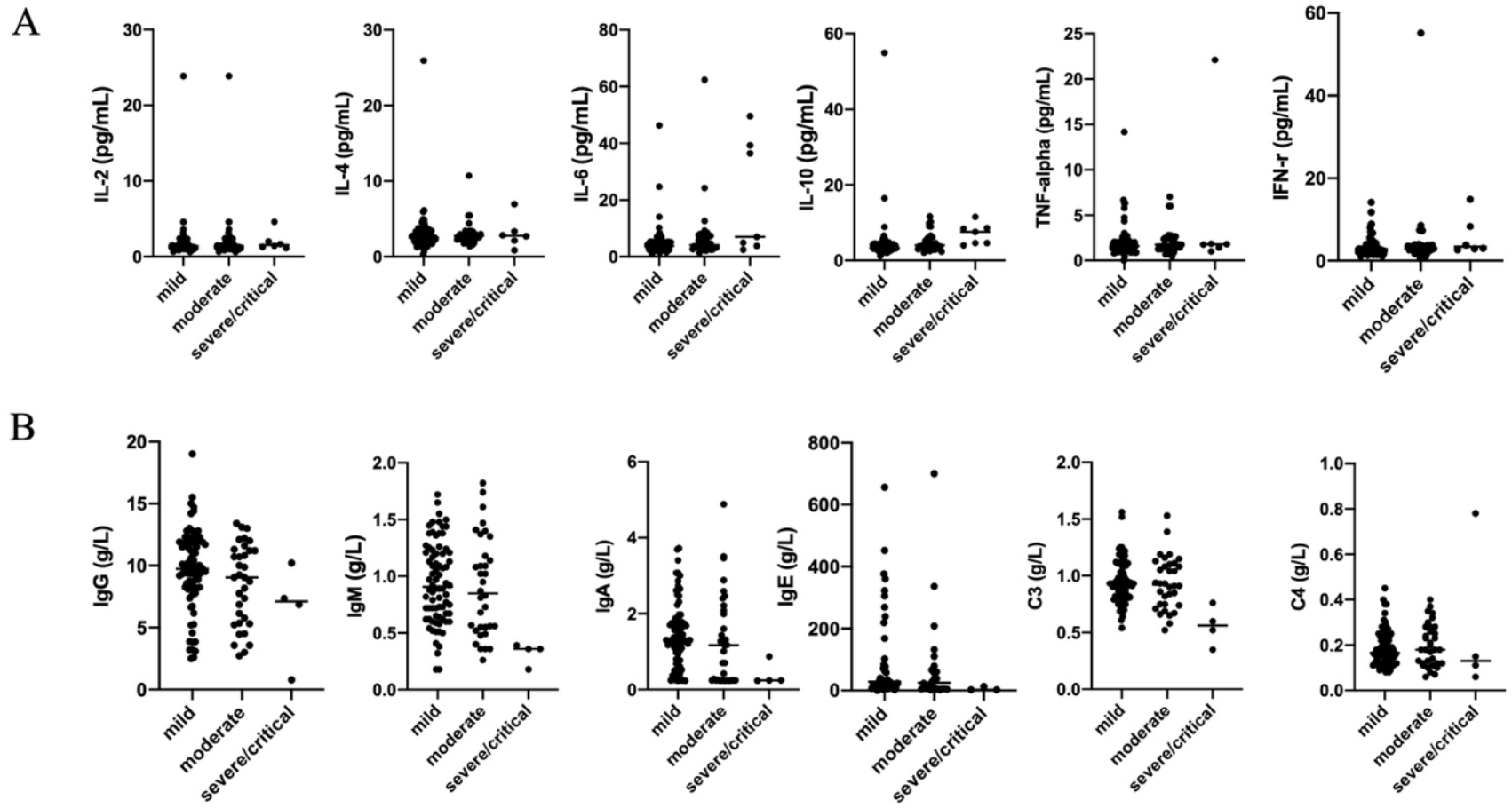
Levels of cytokines, immunoglobulins and complement in children with COVID-19 patients. A. Cytokines (IL-2, IL-4, IL-6, IL-10, TNF-*α*, and IFN-*γ*) in patients in different groups; B. Immunoglobulins (IgG, IgA, IgM, and IgE) and complement (C3 and C4) in different groups.

**Figure 2.**
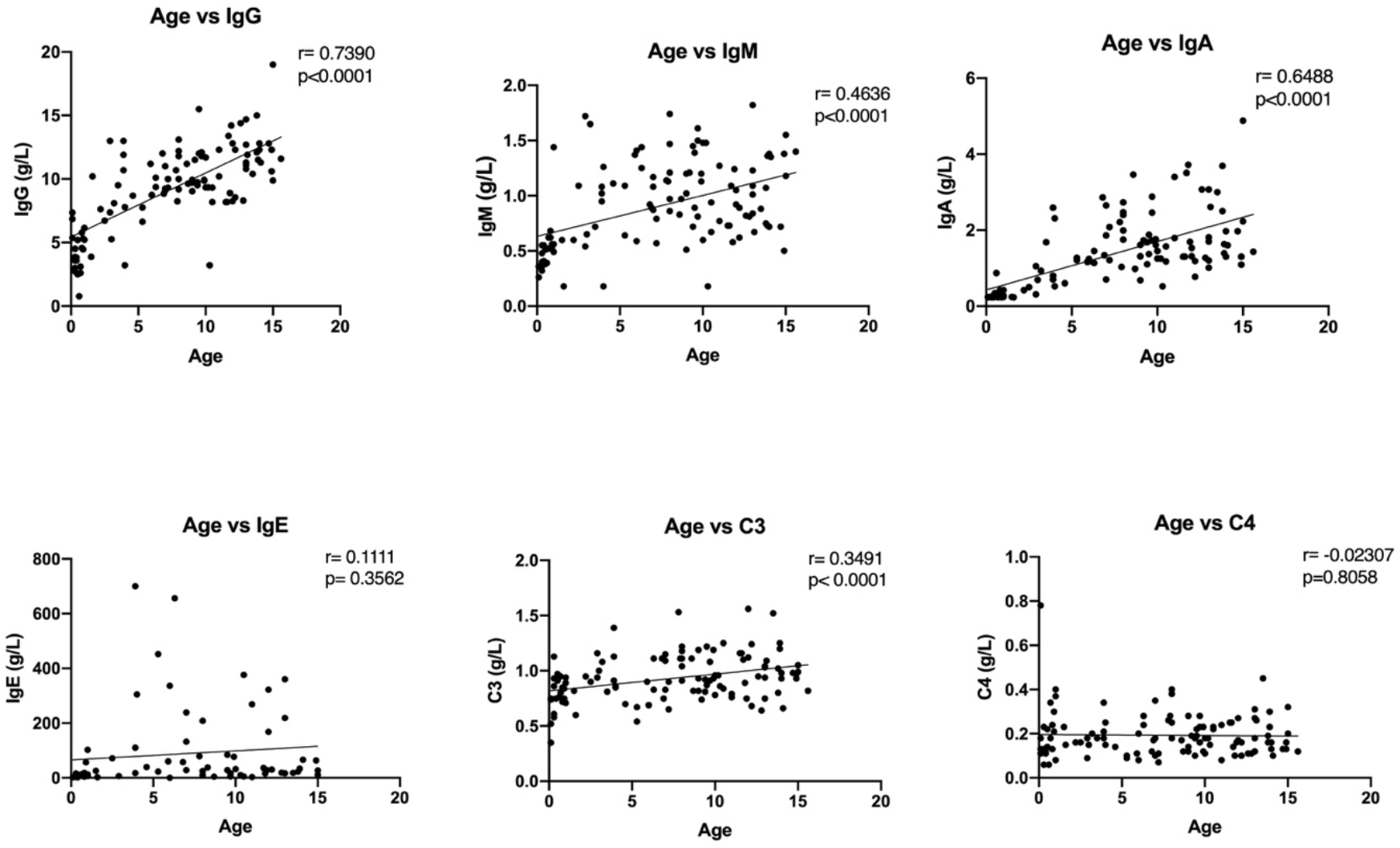
Levels of immunoglobulins (IgG, IgA, IgM, IgE) and complement (C3, C4) relative to age in COVID-19 patients. IgG, IgA, IgM and C3 were significantly higher in older children, while IgE and C4 were not associated with age in COVID-19 patients.

### Association of inflammatory markers with disease severity

To analyse potential markers of disease severity, we compared potential markers in mild cases with those with moderate or severe/critical disease. Individual analyses showed significant association (not corrected for multiple testing) between severity and age, CRP, IL6, IL10 and D-dimer levels. However, in a multivariate logistic regression analysis the only independent predictor of severe disease was age (P<0.001).

## Discussions

In this paper we present novel data on the immune responses seen in children hospitalised with COVID-19. A total 127 children with COVID-19 were admitted to hospital. The majority were male (67.7%), and the median age was 7.3 years. Patients with severe disease were younger (only 1 was over the age of 2) and generally had significant co-morbidity. The immune responses are significantly different from adults. Neither numbers of helper T cells and suppressor T cells were associated with severity of disease. Furthermore, our data show that SARS-CoV-2 does not trigger a severe inflammatory response or ‘cytokine storms’ in children, which potentially explains the much better outcomes seen in children infected with SARS-CoV-2. This was despite the presence of viral pneumonia on CT scanning in over half of the children studied.

Lymphopenia is common in adults with COVID-19, but seems to be rare in children. In adults, lymphocyte counts also show negative correlations with the severity of disease and with the viral copy numbers from nasal or throat swab specimens.^14,15^ The subsets of lymphocytes which are reduced in number in adult patients include CD4+ T cells, CD8+ T cells, B cells, NK cells, memory and regulatory T cells.^14,16^ In adults, lymphocyte counts will increase during the convalescent stage of disease. In one recent study, enrichment of genes in apoptosis and P53 signalling pathways from bronchoalveolar lavage fluid (BALF) and peripheral blood mononuclear cell (PBMC) was observed suggesting, SARS-CoV-2 infection probably induces lymphocyte apoptosis.^17^ Our data are in keeping with one previous small case series of 34 children in which only one case presented with lymphopenias.^18^ Two preliminary reports have suggested that in contrast CD4+ T cell count and CD8+ T cell count may increase in children during COVID-19.^19,20^ Our study shows that for children with COVID-19 the numbers of CD4+ T cells, CD8+T cells, B cells, NK cells are not reduced, even in moderate or severe disease. In addition, the cell number of different lymphocyte subsets did not change between measures taken at admission and immediately before discharge.

Importantly, our data are also in keeping with a recent large epidemiology study characterising demographic features in children with COVID19 in China. This study reported that half of the patients with severe disease were less than one year of age.^21^ Our study also shows that the severe/critical cases often have comorbidities which might be an important explanation for case severity amongst children with COVID-19.

Cytokines are important mediators of the inflammatory response to COVID-19 in adults and are associated with disease severity. A number of clinical trials are ongoing in adults assessing anti-cytokine approaches to improve outcomes in COVID19. However, little is known regarding the cytokine response in children with COVID19. We found that the levels of a range of proinflammatory cytokines including IL-2, IL-4, IL-6, TNF-*α* and IFN-*γ* were no different between mild, moderate and severe/critical groups of children with COVID-19. These data strongly suggest that the better outcomes for children with COVID19, even in the presence of significant viral pneumonia on CT scanning, may be because children do not mount the profound inflammatory response to infection with SARS-CoV-2 seen in some adults and are thus less prone to multi-organ damage. In addition, it would seem unlikely that anti-cytokine approaches will be effective in patients in this age group. Recently, a subgroup of children infected with SARS-CoV-2 have been described who go on to develop a severe inflammatory condition which shows some similarities to Kawasaki syndrome, and which has been called Paediatric Inflammatory Multisystem Syndrome Temporally associated with SARS-CoV-2 (PIMS-TS).^11^ These patients do show increased inflammatory markers including CRP and ferritin and some also have elevated markers of myocardial damage: it is conceivable the child in our study who died aged 7 had this condition. However, this appears to be a very rare complication.

This study has some limitations. First, this is a retrospective and observational study using pre-existing electronic medical record data. Second, there are relatively few children with severe disease due to COVID-19 and so we may have missed small differences in cytokine levels or cellular responses between the groups with different disease severity. Finally, we did not have measures of viral load and hence cannot take this into account in our analyses. Nonetheless, we show convincingly that the majority of children with COVID19 admitted to hospital have a good outcome unless they have significant co-morbidities, and that this may in part be due to the failure of younger patients to mount a major cytokine response to the SARS-CoV-2.

## Data Availability

Please send email to Professor Jin Mei

## Contributors

GQ designed, analysed and interpreted the results, and wrote the Article. YZ, YX, WH collected data and analysed the results. IPH interpreted results and critically reviewed the manuscript. JY, HL, LD, and LR designed and analysed the results. MY and JM designed, interpreted the results, and coordinated the study.

## Acknowledgements

IPH is a National Institute for Health Research (NIHR) Senior Investigator.

## Declaration of interests

We declare no competing interests.

## Notes

### Competing Interest Statement

The authors have declared no competing interest.

### Funding Statement

This study was funded by the National Natural Foundation of China (No. 81970653).

### Author Declarations

This study was conducted in accordance with the Declaration of Helsinki and was reviewed and approved by the Medical Ethical Committees (2020-R120).

